# Family satisfaction with involvement in decision making in the intensive care unit: A scoping review

**DOI:** 10.1101/2024.03.11.24304110

**Authors:** Ntombifikile Klaas, Onalenna Baliki

## Abstract

**Background:** Decision making in the intensive care unit (ICU) is often complex, involving frequent interactions between patients, families, and health care professionals. Family members play an important role as surrogate decision markers because patients admitted in ICU lack decision-making capacity due to their critical state. Lack of regular, timeous, up to date and honest information provided by the ICU healthcare professionals may negatively influence the family members’ decision making ability and overall satisfaction.

**Aim:** To identify existing literature that describe family satisfaction with involvement in decision making in the intensive care unit.

**Methods:** A scoping review of literature guided by Arksey and O ‘Malley’s framework was conducted. Literature search was completed using five databases. Primary studies published in English between January 2010 and December 2023 were retrieved and analyzed using thematic analysis.

**Results:** Of the 152 studies identified during the search, 23 were eligible for inclusion. Most of the studies (n = 19; 83%) focused on family satisfaction with care and decision making and four (n=4; 17%) focused on strategies aimed at enhancing family involvement in decision making. These strategies included: testing the effects of using information booklets, structured attendance of interdisciplinary ward rounds, development and implementation of evidence-based communication algorithm and exploring the culture of interprofessional collaboration. The three themes that emerged from the scoping review were: *satisfaction with involvement, communication, and support*.

**Conclusion:** The use of structured communication programs as well as regular, timeous and honest information about the patient’s diagnosis, treatment plan and prognosis are vital measures to enhancing family involvement in decision making.

## Introduction

Admission in the Intensive care unit (ICU) due to critical illness usually comes unexpectedly, leaving the family members of the critically ill patient helpless and confused. It has the potential to disrupt the steady state of internal equilibrium usually maintained in the family unit. It can change the lives of the patient and those close to them in a minute, leaving little time for family adjustment. ^[1]^ Family members of critically ill patients may experience stress, anxiety, depression and post-traumatic stress during admission and after their loved one has been discharged from ICU. The intimidating environment of the ICU, with its complex and technological equipment, adds to the distress and discomfort of family members particularly because they are not psychologically prepared for a member’s critical illness. All these factors alter the family members’ decision making ability.

The findings of a review on involvement of ICU families in decision making reported that 70% of families exhibited symptoms of anxiety and 35% showed signs of depression in the first few days after admission of a family member in ICU. ^[2]^ A Brazilian study on symptoms of anxiety and depression in family members of ICU patients with a sample of 135 family members, found that 28.9% of family members experienced depression whereas 39.3% experienced anxiety. ^[3]^

Family centered care (FCC) framework recognizes that patients form part of a social structure and web of the relationship between the healthcare providers (HCP) and families in the ICU. ^[4]^ Family members play an important role as surrogate decision markers because patients admitted in ICU lack decision-making capacity due to their critical state. ^[4, 5]^ Therefore, family members are responsible for making decisions like consenting for a procedure, treatment, care as well as end of life decision for the patient such as withholding or withdrawing therapy. ^[6]^

Families of critically ill patients admitted in ICU remain an integral part of the healthcare system. Reeves et al. ^[7]^ emphasizes that families have a unique contribution towards healing and patient care as they act as advocates for the sedated and intubated patients. Family members play a key role as central hubs of information and the source of support for their relatives since they know the patient better in the light of preferences, cultural and religious affiliations ^[5,7]^.

Several studies have focused mostly on dynamics of care including family needs and coping, end of life decision making, and communication. ^[1, 5, 7, 20, 22]^ Meeting the family member’s basic needs in the ICU such as proximity, honest information; reassurance and support facilitate effective coping mechanisms in family members when dealing with critical illness. ^[8, 9]^ Several studies have centered their research on family involvement during specific events such as end of life decision making, family conferences, family presence in resuscitation and family participation during rounds. ^[1,5,7,20,22]^

In order to be active participants in the decision-making process, family members need timeous, transparent, honest and consistent information about their family member’s condition ^[5, 20]^. Findings from studies on family involvement in the care and decision making, show that family members were highly satisfied with the overall care than their involvement in decision making. ^[3,11,12,23]^ Therefore, this study intended to address the gap in literature by exploring family satisfaction with involvement in decision making in the intensive care unit. Comprehensive knowledge regarding the variables impacting family satisfaction may assist healthcare providers in the improvement of the experience of ICU family members. It may also contribute to the early identification and support of family members at risk of a poor ICU experience, which ultimately could be reflected on low satisfaction levels of family members.

## Materials and methods

A scoping review is an exploratory, non-experimental research design that systematically maps available literature and determines the scope or coverage of a body of literature on a given topic, identifying key concepts and gaps in the research.^[15]^ Arksey and O’Malley’s ^[16]^ five-stage framework underpinned the approach for this scoping review. These include: identifying the research question, identifying relevant studies, selection of studies, charting the data, collating, summarizing and reporting the results using the PRISMA flow diagram ^[17]^ The reviewers did not develop a priori protocol.

### Identifying the research question

The aim of this review was to identify existing literature on family satisfaction with involvement in decision making in the intensive care unit. To achieve this aim, the following research question guided the scoping review: ***What is currently known on family satisfaction with the involvement in decision making in the intensive care unit?***

The “**PCC**” acronym was used as a guide to ensure a focused research question. In this study, the mnemonic was denoted: **Population**: Family members; **Concept**: Satisfaction with involvement in decision making; **Context**: Intensive Care Unit.

### Identifying relevant studies

The search strategy involved five electronic databases searched namely: PubMed, CINAHL, MEDLINE, SCOPUS and Academic search. The search terms used included the following: “family satisfaction AND involvement OR participation AND decision making AND adult intensive care unit OR adult critical care unit OR ICU AND family OR relatives OR next of kin.” These search terms were adapted and used for each database. The search strategy was developed with guidance from the librarian.

In this review, primary research studies (qualitative, quantitative and mixed methods) published in English between January 2010 and December 2023 on family satisfaction with involvement in decision making in ICU were considered. Secondary studies such as conference abstracts, conference notes, other reviews, dissertation and thesis were excluded.

A three-step search strategy was used in this review ^[18]^. An initial literature search was conducted using PubMed database to identify the search words appropriate for an exhaustive data search (**Table 1**). This was followed by a comprehensive search of the four selected databases, as well as a hand search of specific intensive care journals. Thirdly, the reference lists of retrieved literature was screened for possible additional studies for inclusion.

### Study selection

The selection of studies obtained from the literature search was conducted based on the general selection criteria for inclusion. The studies identified were exported to the Mendeley reference manager software in order to identify duplicates and gather all the publications. In the first stage of selection, one reviewer (OB) designed and conducted the search strategy supported by the second reviewer (NK). The two reviewers independently screened titles, abstracts, and full-text articles. A total of 48 full text articles were considered relevant and read by the two reviewers independently. The inclusion and exclusion criteria guided this process with 25 studies excluded. A total of 23 studies were included in this review. The PRISMA flow diagram ^[19]^ was used to report and map the search process (**Fig. 1**).

**Fig. 1:**
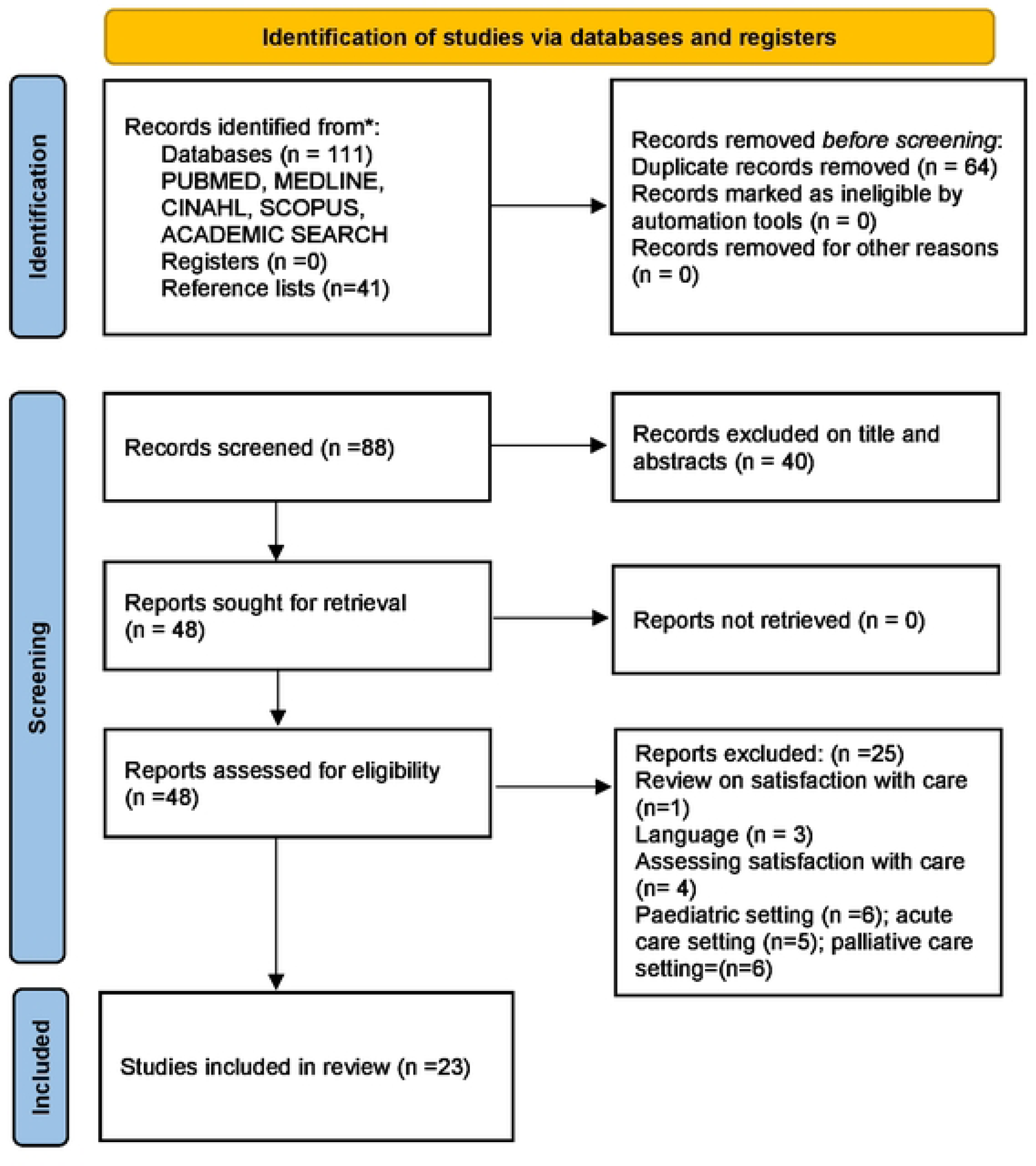
PRISMA Flow diagram of the literature search.

### Charting of data, collating, summarizing and reporting the results

The charting of the results was done using a data extraction sheet adopted from the Joanna Briggs institute. ^[18]^ To enable a logical and descriptive summary of the results, data were extracted using the following headings: author/s, year of publication, country, study aim/ objective, population, sample and sampling, data collection method and key findings. **Table 2** summarises the studies included in this review. The first reviewer extracted the data iteratively, while the second reviewer verified the extracted data.

The findings from each study selected were organized and key themes developed following Braun and Clarke’s thematic analysis method^[19]^. Two reviewers independently read the findings repeatedly to thoroughly familiarise themselves with them. The findings were reported in a narrative format after comparing and contrasting the themes generated.

## Ethical considerations

Permission to conduct this review was obtained from the Research and Ethics Committee, Faculty of Health Science, University of Witwatersrand (W-CBP-210115-01).

## Results

The search yielded a total of 152 potential studies, of which 111 were retrieved from the database search and 41 studies from the reference lists. Sixty-four duplicates were removed, and 40 studies were excluded based on the title and abstract. Finally, 48 full-text studies were further assessed for relevance against the inclusion and exclusion criteria; 25 studies were excluded based on 1 study being a review which focused on family satisfaction with care, 3 studies were conducted in other languages, 4 studies assessed satisfaction with care only, 6 studies were conducted in a paediatric setting, 6 in a palliative care setting and 5 in an acute care setting, leaving 23 studies for inclusion.

Of the 23 studies included in this review, 18 were quantitative, 4 qualitative and 1 mixed methods. The quantitative studies predominantly used study designs such as prospective cohort and descriptive cross sectional with few articles using retrospective cohort, observational and pre-post study design. The qualitative studies used descriptive exploratory and ethnography study designs. The mixed methods study included cross sectional and in-depth interviews.

Most of the studies (n = 19; 83%) focused on family satisfaction with care and decision making and four (n=4; 17%) focused on strategies aimed at enhancing family involvement in decision making. These strategies included: testing the effects of using information booklets, structured attendance of interdisciplinary ward rounds, development and implementation of evidence-based communication algorithm and exploring the culture of interprofessional collaboration.

The studies included in the review presented data from five different continents representing 15 countries with diverse cultural backgrounds. North America was the leading continent with 9 studies, United States of America 4, Massachusetts 2, Canada 2, UK and Canada 1. The second continent was Europe with 6 studies (Norway 3, UK 1, Germany 1, Denmark and Netherlands 1. The third continent was Asia with 5 studies. Malaysia, 2, Bahrain 1), Hong Kong 1 and India had 1 study. Australia had one study and the last two studies were from Africa (Kenya 1 and South Africa 1).

Participants included in most of the studies (n=21; 91%) were family members of critically ill patients admitted in ICU and two (n= 2; 9 %) included family members and HCPs. The participants of each study ranged from six and 7173 family members and 9 to 56 HCPs. Three key themes were identified from the scoping review: *satisfaction with involvement, communication and support.* **Table 3** summarizes these themes.

### Theme One: Satisfaction with involvement

Satisfaction of family members with involvement was the most investigated theme in this review. Two key areas related to satisfaction with involvement were identified, namely: satisfaction with involvement in decision making and satisfaction with involvement with care.

#### Satisfaction with involvement in decision making

Family satisfaction with involvement in decision making was presented by nineteen several authors in this review. ^[4,5,7,10, 20, 21, 22, 24,14,23,26,27,29,30, 31, 32, 33, 34,40,41]^ It was found that amongst the quantitative studies, the family satisfaction in the intensive care (FS-ICU 24) survey, a 24 item self-report that measures family satisfaction with care and decision making was used. Family members rated decision making domain with the mean score between 64.8 and 80.6. ^[5,7,10, 14, 20,21, 22, 24,14,23,26,27,28, 29,30, 31, 32, 33, 35, 36, 37]^

The quantitative studies examined inclusion in decision making through identifying and describing the inclusion of family members in the decision making in the ICU and comparing perspectives of family members of non-survivors and survivors. ^[5,25, 26,27, 29]^ They found that family members of non-survivors were more satisfied with inclusion in decision-making than families of ICU survivors. Family members perceived that they had not been involved in decision making to start life support intervention, however, most frequently involved in decision making just before decisions to withhold or withdraw life support and provide comfort care intervention. ^[7,20,22]^

Poor communication and lack of support were highlighted as the two major barriers that led to dissatisfaction with family involvement in decision making. Family members expressed dissatisfaction with communication during the decision-making process in the ICU; they felt that there is a need for transparent, honest and consistent information. ^[5, 20]^ The findings of the Malaysian ^[29]^ study aimed at testing the effects of information booklets on family satisfaction with decision making revealed higher scores of satisfaction after the intervention. This indicates that the structured communication program improved family members’ experiences on decision making.

This was evident in the Norwegian study, with the population of 123 relatives found that family members of a patient who died during the stay were more satisfied with support in decision making compared to family members of ICU survivors. ^[5]^

#### Satisfaction with involvement in care

The focus of this review was to gather evidence on studies which looked at family satisfaction with involvement in decision making, however, this subtheme emerged because the FS-ICU 24 survey assesses satisfaction with care and decision making. ^[5,7,10, 14, 20, 21, 22, 24,14,23,26,27,29,30, 31, 32, 33, 35, 36,40,41,42]^

This review revealed that family members were not entirely satisfied with care rendered to their loved ones in the intensive care unit. Some of the factors that emerged under this sub theme contributed to lower scores. These factors include dissatisfaction with pain management, agitation management, waiting room’ s atmosphere, emotional support and communication ^[23,24]^. Family members reported that they desired to be involved or participate in the direct care of their loved one, however, the critical care nurses were reluctant to involve families in the care. ^[20, 22]^

Evidence shows that involving families in patient care helped family members maintain the family bonds and cohesiveness. It also helped minimize the disturbance imposed on the family unit following a family member’s admission to the ICU. ^[20, 21]^

Family members in this review were willing to be involved in their patient care and they believe that the patients could still hear and feel their presence even if they were not able to see or talk ^[20,22]^ Involving family members in direct patient care may pose some challenges on the family members such as fear of doing such activities with their ill relative surrounded by machines, attached to so many lines and tubes without the nurse to direct them. ^[22]^

### Theme Two: Communication

Communication emerged as the second theme in this review. The three sub themes that emerged were: consistency of information, clarity and completeness of information and transparency.

#### Consistency of information

This review revealed that the information need was perceived important by the family members in the ICU, hence they reported less satisfied with the consistency of information they received about their patient ^[5, 9, 22]^. A prospective cohort study ^[31]^ conducted in Massachusetts with 124 participants found that family members reported one or multiple episodes of inconsistent information during their relatives’ admissions.

It was also found that poor or inconsistent communication between the ICU health care professionals made it difficult for family members to obtain consistent information about their relatives. ^[9]^ In this review, family members were not satisfied with frequency of communication with physicians; their perception were that it is difficult to be addressed by the doctor. ^[27, 31]^

#### Clarity and completeness of information

The second subtheme that emerged from communication is clarity and completeness of information received by family members in the ICU. Fateel and O’ Neil ^[21]^ conducted a study in Ireland and found out that most family members of the critically ill patient in the ICU are not satisfied with the communication. They felt that they did not receive full information about the patient which might hinder their participation in the decision-making process. In addition, a Canadian study by Henrich *et al.* ^[37]^ revealed that family members believe that they were able to handle the honest information about their patient and that full and realistic information about the situation enables them to make better treatment and end-of-life decisions and to cope with the patient’s situation. A Kenyan study ^[38]^ on family involvement in the provision of care to critically ill patients revealed that family members were not satisfied with the depth of information about the patient care, and families reported build-up of anxiety levels due to withholding of information by nurses.

#### Transparency

The third sub theme was transparency in communication. Family members were not allowed to be around during the doctor’s rounds, and these left them with insufficient information. ^[21, 22]^. Family members expressed the need for transparency therefore they desire to know exactly what going on with their patient and these may enhance the ability of the family to support and protect the patient through decision making process. ^[20]^

### Theme Three: Support

The third theme that emerged was support. Informational and emotional support were identified as subthemes. Thirteen studies sought to identify and explore the support given to family members during the clinical decision-making process. ^[4,5,20,21, 22,25, 27,28, 29, 33–35,40]^

#### Informational support

Support with information emerged as an important need for family members caring for a relative in the ICU. It is the responsibility of health care providers (doctors and nurses) in the ICU to ensure that this need in met. This review revealed that most family members were not satisfied with the information given about their patient; they reported that nurses withhold information from them as nurses could not share some information. ^[22,36,40]^

#### Emotional support

Emotional support emerged as the second subtheme under support. Family members expressed some form of distress due to the critical illness of their loved one as they were not prepared for it. Therefore, they desired to be emotionally supported particularly by the nurses. ^[21,22,42]^ In this review, family members reported the need to be reassured about the comfort care given to the patient and the nurses to be patient with them during this critical time. ^[21]^

## Discussion

To the best of our knowledge, this is the first scoping review to describe published literature on family satisfaction with involvement in decision making in the intensive care unit. The review shows that the level of family satisfaction with involvement in decision-making can be improved when the HCPs put in place some strategies to facilitate family participation in the decision-making process. ^[4]^ Communication and receiving information in the ICU emerged as an area that needs improvement because family members felt that it hinders their participation in the decision-making process. ^[9.21]^ These results are consistent with the results of an Australian study ^[37]^ where family members were not satisfied with the frequency of communication with both ICU nurses and doctors. Family members should receive clear, timeous, honest and consistent information to enhance shared decision-making process. ^[37]^

The scoping review findings have demonstrated that evidenced-based structured communication strategies are effective to improve family members’ satisfaction with participation in decision making. ^[5,28]^

An evidence-based structured communication algorithm was used to identify clinical triggers for family meetings in a United States of America study ^[4]^. There were statistically significant improvements noted for how often the family members were able to participate in decision making where before the algorithm, the score was 45% and after the algorithm was 68%. ^[4]^ The Malaysian study by Othman et al. ^[30]^ investigated the effects of information booklets on family members’ satisfaction with decision making in ICU with the population of 84 family members. There was improvement in the FS-ICU decision making mean score pre-test score of 22.7 compared to the post-test score of 29.6 ^[30]^.

A retrospective cohort study conducted in the USA with a population of 453 families found that the families who reported attending the family conference were more satisfied with the decision-making process in the ICU ^[7]^. This confirms that the use structured communication program improves family satisfaction with decision making. In contrast, in the study conducted in the USA by Jacobowski et al. ^[36]^ where family rounds were implemented to improve communication in the ICU found that family members reported that they were not satisfied with the time given to address concerns and questions answered during decision making process before and after attending family rounds.

Another theme that was highlighted in this scoping review was involvement during decision-making process. Even though family members assume surrogate decision-maker roles over the decision-making process about the choices of the diagnostics, treatment, and life support care of their loved one, they have expressed dissatisfaction when it comes to the timing of inclusion during decision making. The participants in a Canadian study by Kryworuchko et al. ^[6]^ expressed that they were not involved in decision making to start mechanical ventilation or other interventions. However, they were most frequently involved in decision making just prior to decisions to withhold or withdraw life support and provide comfort care interventions. This is confirmed in the Norwegian study by Haave et al. ^[41]^ with the population of 57 family members where degree of satisfaction was found to be lower in relation of family members participation in the decision making process. Furthermore, the Bahraian study by Hamed et al. ^[42]^ found similar results where satisfaction with care scored higher than satisfaction with decision making.

Family involvement during the decision-making process particularly in end-of-life decision to withhold or withdraw life support made for critically ill patients was found to vary from active participation in the decision making process to acceptance of the physicians’ decision or just receiving information of the physicians’ decision. ^[20]^ In a situation where the family members were just informed about the physician’s decision to terminate treatment, families were unsure whether the patient was competent to decide considering the critically ill state of the patient and fluctuating level of consciousness. ^[20]^ Similarly, in a South African study by Kisorio and Langley ^[24]^ family members were told about the decisions made without being actively involved.

Family members of non-survivors were found to be more satisfied with inclusion in the decision-making process more than with family members of survivors. ^[5, 25]^ This can be attributed to the fact that family members of non-survivors are often involved in decision-making just before withholding or withdrawing life support and provide comfort care interventions. ^[7]^ Also, the difference may be explained by the fact that some patients who are admitted in ICU can participate in the decisions and consent for procedures for themselves. ^[5]^

This review also highlighted the importance of support from HCPs during the care and decision-making process which is crucial for the well-being of family members and their ability to participate in making decisions. Family members require informational support as well as emotional support to assist them to cope with the critical illness of their loved ones. The information need of the family members remain unmet. The family members were not satisfied with the consistency of information about the patient’s condition and the completeness of the information. ^[5,7,21]^ A South African study ^[24]^ with a population of 17 family members found similar findings that family members were not satisfied with the information they received about their patient’s progress.

In a qualitative study done in Ireland with a sample of six family members of a critically ill patient, family members highlighted that the nurses should have more patience when dealing with them during this critical time and support them emotionally. ^[1]^ It is also emphasized that the ICU health care providers to have a thorough consideration of emotional impacts imposed by the admission of a family member in the ICU so that they may understand the family needs to be holistically supported. ^[1]^ Findings from the study by Kisorio and Langley ^[24]^ concurs that those family members of critically ill patients need emotional support to cope, however, family members reported dissatisfaction with emotional support in the ICU. The importance of emotional support was found to be a strategy that can be adopted by ICU health care providers to increase the satisfaction of the family members. ^[42]^

A quantitative study done with a population of 215 in Germany found compassion and emotional support to be important particularly during the decision-making process and recommends that a shared decision-making model for family participation has to be adapted to family needs. ^[33]^ The findings of a Swedish study by Blom et al. ^[40]^ confirmed that support in the form of information sharing and emotional support is important for family members to enable them to participate in the decision making.

## Limitations of the study

The following limitations identified in this review are worth noting:

Due to the date limit and language restriction, we may have missed important studies published before and after the set dates and others published in other languages. There were only five databases used, which may have limited the search.

## Recommendations

The findings from this review suggest that satisfaction with family involvement in decision making in ICU has not been investigated thoroughly within the critical care literature. Research gaps identified in this review suggest that extensive research is still needed to be conducted on the nature and extent of family involvement in decision making process in the intensive care unit. Future research is required also to investigate the factors that influence family involvement in decision making in the ICU including socio-cultural, organizational and contextual factors.

Future research should consider triangulation of methods when investigating satisfaction with family involvement in decision making in the ICU to improve and expand the findings. The future research might be strengthened by incorporating the ethnographic approaches that may produce in depth and context specific findings for family satisfaction with involvement in decision making. In this scoping review, findings indicate that more studies have been done globally but in Africa there is lack of similar research. We recommend more research to be conducted in Sub-Saharan Africa focusing on family satisfaction with involvement in decision making in ICU considering the unique racial, cultural, ethnic, and linguistic differences found in African countries.

## Conclusion

Despite the inclusion of studies from fifteen countries with diverse cultural, religious and health policies and regulations, family members were mostly dissatisfied with their involvement in decision making. The key finding that emerged from this scoping review is that family satisfaction with involvement in decision making in ICU has not been thoroughly explored.

## Data Availability

All relevant data are within the manuscript and its Supporting Information files.

## Acknowledgements

The authors wish to acknowledge the University of the Witwatersrand for making their library available for the data search needed for this scoping review.

## References

1. Morton, P. & Fontaine, D. (2013). Critical care nursing: A holistic approach. 10th ed. USA: Wolters Kluwer Health Lippincott Williams and Wilkins.

2. Azoulay, E., Chaize, M. & Kentish-Barnes, N. (2014). Involvement of ICU families in decisions: fine-tuning the partnership. Annals of Intensive Care, 4(37), 1–10

3. Fonseca, G. M., Freitas, K. S., Silva Filho, A. M., et al. (2019). Anxiety and depression in family members of people hospitalized in an intensive care unit. Psicologia: Teoria e Prática, 21(1), 328–343.

4. Huffines, M., Johnson, K.L., Naranjo, L.L. S., et al. (2013). Improving Family Satisfaction and Participation in Decision Making in an Intensive Care Unit. Critical Care Nurse 33 (5), 56–68.

5. Frivold, G., Slettebo, A., Heyland, D.K., et al. (2018). Family members satisfaction with care and decision making in intensive care unit and post –stay follow up needs-a cross sectional survey study. Nursing open, 5(1), 6–14.

6. Kryworuchko, J., Stacey, D., Peterson, W.E., et al. (2012). A Qualitative Study of Family Involvement in Decisions about Life Support in the Intensive Care Unit. American Journal of Hospice & Palliative Medicine, 29(1), 36–46.

7. Reeves, S., Sarah E. McMillan, S.E., et al. (2015). Interprofessional collaboration and family member involvement in intensive care units: emerging themes from a multi-sited ethnography. Journal of Interproffesional Care. 29(3), 230–237

8. Nolen, K.B. & Warren, N.A. (2014) Meeting the Needs of Family Members of ICU Patients. Critical Care Nursing, 37(4), 393–406

9. De Beer, J. & Brysiewicz, P. (2016). The needs of family members of Intensive Care Unit Patients: A grounded theory study. Southern African Journal of Critical Care, 32(2), 44–49.

10. Kodali, S., Stametz, R.A., Bengier, A.C., et al. (2014). Family experience with intensive care unit care: Association of self-reported family conferences and family satisfaction. Journal of Critical Care, 29, 641–644.

11. Kleinpell, R., Zimmerman, J., Vermoch, K. L., et al. (2019).Promoting Family Engagement in the ICU: Experience From a National Collaborative of 63 ICUs. Critical Care Medicine, 47 (12), 1692–1698

12. Kohi T.W., Obongo M.W. & Mselle L.T. (2016). Perceived needs and level of satisfaction with Care by far members of critically ill patient at Muhimbili National Hospital intensive Care units, Tanzania. Biomedicice Central, 15, 18.

13. Mujumdar, M.R. & Bhole, S. (2018). To evaluate the level of Satisfaction in Relatives of intensive Care units Patient. National Journal of integrated Research in Medicine, 9(1), 103–107.

14. Clark, K., Milner, K.A., Beck, M., et al. (2016). Measuring Family Satisfaction with Care delivered in the intensive care unit. Critical Care Nurse, 36(6), 08–14.

15. Tricco AC, Lillie E, Zarin W, O’Brien KK, Colquhoun H, Levac D, et al. PRISMA extension for scoping reviews (PRISMA-ScR): checklist and explanation. Annals of internal medicine. 2018;169(7):467–73. pmid:30178033.

16. Arksey H, O’Malley L. (2005). Scoping studies: towards a methodological framework. International Journal of Social Research Methodology, 8, 19–32.

17. Aromataris, E. & Munn, Z. (Editors). (2017). Joanna Briggs Institute Reviewer’s Manual. The Joanna Briggs Institute, Available from https://reviewersmanual.joannabriggs.org/.

18. Peters, M., Godfrey, C., Mclnerney, P. (2021) The Joanna Briggs Institute Reviewers’ Manuel: Methodology for JBI Scoping Reviews. The Joanna Briggs Institute.

19. Braun, V. & Clarke, V. (2006). Using thematic analysis in psychology. Qualitative Research in Psychology, 3(2), 77–101.

20. Hansen L., Rosencrantz S.J., Mularski R.A., et al. (2016). Family perspective on overall care in the Intensive care. Nursing Research, 65 (6), 446–454.

21. Fateel E.E & O’Neill C.S (2016). Family members’ involvement in the care of critically ill patients in two intensive care units in an acute hospital in Bahrain: The experiences and perspectives of family members’ and nurses’ - A qualitative study. Clinical Nursing Studies, 4(1), 57–69.

22. Lind, R., Nortvedt, P., Lorem, G., et al. (2012). Family involvement in the end-of-life decisions of competent intensive care patients. Nursing Ethics, 20(1), 61–71.

23. Mitchell, M.L. & Wendy Chaboyer, W. (2010). Family Centred Care-A way to connect patients, families and nurses in critical care: A qualitative study using telephone interviews. Intensive and Critical Care Nursing. 26, 154–160.

24. Kisorio & Langley. (2016). End-of-life care in intensive care unit: Family experiences. Intensive and Critical Care Nursing, 35, 57–65

25. JanardhanIyengar, S.M., Srinivasan, R., Venkateshmurthy, B.M., et al. (2019). Family Satisfaction in a Medical College Multidisciplinary Intensive Care Unit (ICU)—How Can We Improve? Indian Journal of Critical Care Medicine, 23(2), 83–88.

26. Lam, S.M., So, H.M., Fok, S.K., et al. (2015). Intensive care unit family satisfaction survey. Hong Kong Medicine Journal, 21,435–43

27. Hwang, D.Y., Yagoda, D. Perrey, H., et al. (2014). Assessment of Satisfaction with Care Among Family Members of Survivors in a Neuroscience Intensive Care Unit. Journal of Neuroscience Nursing, 46(2), 106–116

28. Ferrando, P., Gould, D.W., Walmsley, E., et al. (2019) Family satisfaction with critical care in the UK: a multicentre cohort study. Biomedical journal Open, 9, 1–9

29. Maxim, T., Alvarez, A., Hojberg, Y., et al. (2019). Family satisfaction in the trauma and surgical intensive care unit: another important quality measure. Trauma Surgical Acute Care Open, 4, 1–5

30. Othman, H., Subramanian, P., Ali, N.A., et al. (2016). The Effect of Information Booklets on Family Members’ Satisfaction with Decision Making in an Intensive Care Unit of Malaysia. Journal of Young Pharmacists, 8, (2), 128–132.

31. Jensen, H.I., Gerritsen, R.T., Koopmans, M., et al. (2017). Satisfaction with quality of ICU care for patients and families: the euroQ2 project. Critical Care 21:239, 1–10.

32. Hagerty, T.A., Velázquez, A., Schmidt, J.M., et al. (2016). Assessment of satisfaction with care and decision-making among English and Spanish-speaking family members of neuroscience ICU patients. Applied Nursing Research, 29, 262–267.

33. Hwang, D.Y., Yagoda, D., Perrey, H.M., et al. (2014). Consistency of communication among intensive care unit staff as perceived by family members of patients surviving to discharge. Journal of Critical Care, 29, 134–138

34. Sundararajan, K., Sullivan, T.R. & Chapman, M. (2012). Determinants of family satisfaction in the intensive care unit. Anesthetist Intensive Care, 40, 159–165.

35. Schwarzkopf D, Behrend S, Skupin H., et al. (2013). Family satisfaction in the intensive care unit: a quantitative and qualitative analysis. Intensive Care Medicine, 39, 1071–1079

36. Jacobowski N. L., Girard T. D., & Mulder J.A. et al. (2010). Communication in critical care: family rounds in the intensive care unit. American Journal of critical Care, 19 (5), 421–430.

37. Henrich, N. J., Dodek,P., Heyland, D., et al.(2011) Qualitative analysis of an intensive care unit family satisfaction survey. Critical care medicine, 39(5), 1000–1005.

38. Maina P.M, Kimani S & Omuga B. (2018). Involvement of Patients’ Families in Care of Critically Ill Patients at Kenyatta National Hospital Critical Care Units. American Journal of Nursing Science, 7(1), 31–38.

39. McLennan, M. & Aggar, C. (2020), Family satisfaction with care in the intensive care unit: A regional Australian perspective. Australian Critical Care, 33, 518–525.

40. Blom H., Gustavsson C. & Annelie Johansson Sundler A. J. (2013) Participation and support in intensive care as experienced by close relatives of patients-A phenomenological study. Intensive and Critical Care Nursing, 29, 1–8.

41. Haave R.O.,Bakke H.H & Schoroder A.(2021). Family satisfaction in the intensive care unit, a crosssectional study from Norway. BMC Emergence medicine.21:20

42. Hamed KA, Buzaid F, AlHafi M, et al. A cross-sectional study assessing family satisfaction in the intensive care environment in Bahrain: opportunities for improvement [version 1; peer review: 1 approved with reservations] F1000Research 2023, 12:325 10.12688/f1000research.128264.1

43. Rahman W.N.AW.A., Othman A.K.I., Yusop Y.M., et al(2021). Developing family Satisfaction with care in Adult critical care public Hospital Terengganu, Malaysia. The Journal of theory and Practice.2 :(2) 31–35

